# End-to-End Protocol for the Detection of SARS-CoV-2 from Built Environments

**DOI:** 10.1101/2020.08.16.20172668

**Authors:** Ceth W. Parker, Nitin Singh, Scott Tighe, Adriana Blachowicz, Jason M. Wood, Arman Seuylemezian, Parag Vaishampayan, Camilla Urbaniak, Ryan Hendrickson, Pheobe Laaguiby, Kevin Clark, Brian G. Clement, Niamh B. O’Hara, Mara Couto-Rodriguez, Daniela Bezdan, Chris Mason, Kasthuri Venkateswaran

## Abstract

Severe acute respiratory syndrome coronavirus 2 (SARS-CoV-2), the virus that causes coronavirus disease 2019, is a respiratory virus primarily transmitted from person to person through inhalation of droplets or aerosols, laden with viral particles. However, as some studies have shown, virions can remain infectious for up to 72 hours on surfaces, which can lead to transmission through contact. For this reason, a comprehensive study was conducted to determine the efficiency of protocols to recover SARS-CoV-2 from surfaces in built environments. This end-to-end (E2E) study showed that the effective combination of monitoring SARS-CoV-2 on surfaces include using an Isohelix swab as a collection tool, DNA/RNA Shield as a preservative, an automated system for RNA extraction, and reverse transcriptase quantitative polymerase chain reaction (RT-qPCR) as the detection assay. Using this E2E approach, this study showed that, in some cases, SARS-CoV-2 viral standards were still recovered from surfaces as detected by RT-qPCR for as long as eight days even after bleach treatment. Additionally, debris associated with specific built environment surfaces appeared to negatively impact the recovery of RNA, with Amerstat inhibition as high as 90% when challenged with an inactivated viral control. Overall, it was determined that this E2E protocol required a minimum of 1,000 viral particles per 25 cm^2^ to successfully detect virus from test surfaces. When this method was employed to evaluate 368 samples collected from various built environmental surfaces, all samples tested negative, indicating that the surfaces were either void of virus or below the detection limit of the assay.

**Importance:** The ongoing severe acute respiratory syndrome coronavirus 2 (SARS-CoV-2) (the virus responsible for coronavirus disease 2019; COVID-19) pandemic has led to a global slow down with far reaching financial and social impacts. The SARS-CoV-2 respiratory virus is primarily transmitted from person to person through inhalation of infected droplets or aerosols. However, some studies have shown virions can remain infectious on surfaces for days, and can lead to human infection from contact with infected surfaces. Thus, a comprehensive study was conducted to determine the efficiency of protocols to recover SARS-CoV-2 from surfaces in built environments. This end-to-end study showed that the effective combination of monitoring SARS-CoV-2 on surfaces required a minimum of 1,000 viral particles per 25 cm^2^ to successfully detect virus from surfaces. This comprehensive study can provide valuable information regarding surface monitoring of various materials as well as the capacity to retain viral RNA and allow for effective disinfection.

## Introduction

The ongoing coronavirus disease 2019 (COVID-19) pandemic is caused by severe acute respiratory syndrome coronavirus 2 (SARS-CoV-2) (1), which was first identified in Wuhan, China, in December 2019. The World Health Organization (WHO) declared it a Public Health Emergency of international concern on January 30, 2020, and then a pandemic on March 11, 2020. The high infection rate and rapid spread has caused global, social, and economic disruption (2), including postponement of many sporting, religious, political, and cultural events, as well as the closure of non-essential business, schools and universities worldwide across 160 countries (3).

A primary goal set forth by the government has been to keep essential businesses (grocery stores, hospitals, gas stations, etc.) open while protecting staff and patrons with as little disruption as possible given the severity of the situation. Since the current model suggests that the main route of infection is person to person through inhalation of aerosolized droplets containing the virus (4), the use of masks, maintaining physical distancing, avoiding touching ones face, and washing hands have all been identified as important factors in preventing transmission (5). However, since SARS-CoV-2 can remain infective for hours to days on surfaces (6), it is possible to transmit and contract the virus by coming in contact with contaminated surfaces (7). When infected individuals inadvertently carry SARS-CoV-2 into built environments, the infection may spread between individuals via fomites. compromising the ability of workers to continue normal operations and activities. Therefore, disinfection and cleaning regiments have been established by most organizations as a precautionary measure to safeguard against viral transmission (8).

Since SARS-CoV-2 is fatal and a worldwide concern, the Centers for Disease Control and Prevention (CDC) and National Institutes of Health (NIH) have issued directives that molecular (RNA)-based detection be applied to clinical specimens (4, 5) but no such policies have been set for environmental monitoring (6). Since the risk of infection from contaminated surfaces is of serious concern, the need for environmental surface monitoring, along with understanding the effectiveness of cleaning and disinfection is critical. In this study we outline a comprehensive approach to characterize and develop an effective environmental monitoring plan that can be used to understand viral persistence and elimination.

The ability to collect and analyze samples is fundamental to any microbial monitoring analysis. During this study, a noninfectious and replication-deficient virus was used as a surrogate for the SARS-Cov-2 virus to inoculate representative test surfaces and analyzed for recovery efficiency. Several sampling strategies were evaluated for collecting samples from various materials. Experimental parameters such as the method of viral inoculation of each surface type, collection and transport, and analysis techniques were used to determine viral recovery efficiency, total biomass, species-specific recovery, background contaminant levels, inhibitory factors, as well as sampling and detection anomalies.

The overall objective of the study was to develop a standardized end-to-end (E2E) protocol for the detection of SARS-CoV-2 from built environmental surfaces and to determine the minimum number of RNA copies needed on fomites to positively detect virus within the limit of detection of our assay. This study included collecting ~400 samples from seven surface types common to materials found in the built environment and measuring the recovery efficiency of the surrogate SARS-CoV-2 virus. After establishing the E2E protocol, further reproducibility studies were conducted by a second laboratory for verification.

## Results

### Efficiency and Influence of Swab and DNA/RNA/Shield (DRS) Solution on Viral Extraction

To determine the impact of the swabs and DRS transfer medium on the percent recovery of viral particles, the SeraCare AccuPlex SARS-CoV-2 reference material was added to tubes containing water and DRS solution, either directly into the tubes or inoculated onto swabs first, before RNA extraction followed by RT-qPCR. The resulting viral copy numbers were then compared and computed to understand the effects of swabs, DRS solution, and various other combinations in the recovery of viral particles (*Figure 1*). The RNA copies detected from Accuplex placed into the water suspension (no swab) was used as the 100% positive copy number reference to calculate other combinations. For practical applications, swabs should either be placed in water or in a transport medium like DRS so that samples could be transported and processed in the laboratory. Relative to Accuplex in water (no swab), there was a 12% loss of viral load when Accuplex solution soaked on the swab before being placed in water (swab effect). Similarly, when Accuplex was placed directly into DRS instead of water (no swab), the recovery was 71% (DRS effect). The double effect of the swab and DRS on viral recovery was significantly less (p=0.0008), with only 21% recovery (*Figure 1*).

**Figure 1:**
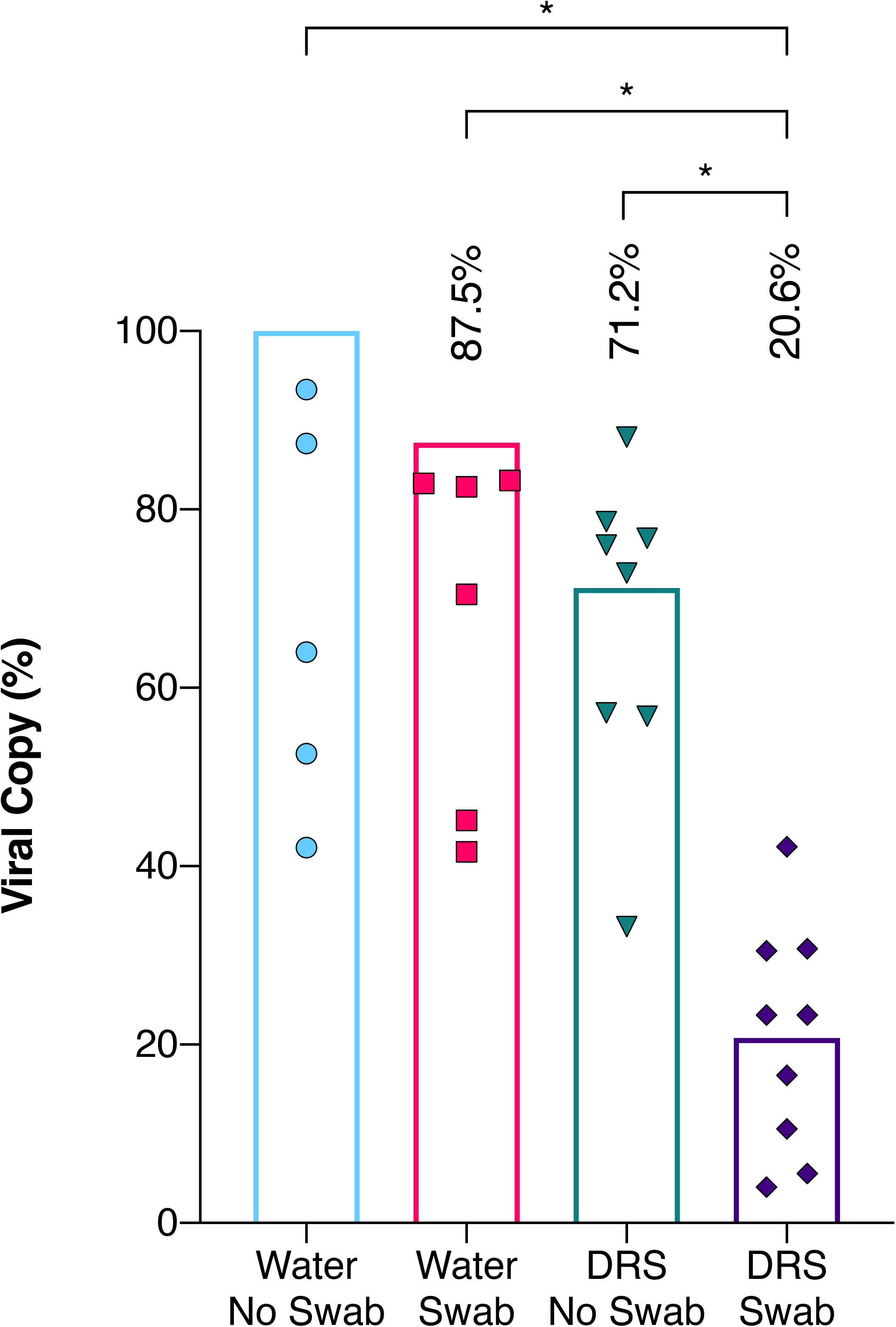
Influence of swab and DRS on viral RNA extraction efficiency. Equal quantities of the inactivated AccuPlex viral particles were extracted using a variety of initial extraction conditions and then quantified using RT-qPCR assay. The extraction conditions encompassed water with no swab (●), water with Isohelix swab (■), DNA/RNA Shield (DRS) with no swab (▼) DRS with Isohelix swab (♦). Each extraction condition was then divided by the average copy numbers generated from the water with no swab (theoretical highest yield) to get percent recovery and plotted with columns representing their mean percentage. Welch’s t test was used to determine significant differences between extraction conditions, significance (p<0.05) denoted by ‘*’.

### RNA Extraction Efficiency

An automated RNA extraction system was compared to a manual extraction where AccuPlex was inoculated on swabs containing DRS (*Figure 2*). There were no significant differences between the automated system and the manual extraction method. Subsequently, several viral transport media were also compared with water. No significant differences were observed between the three different viral transport media (*Figure 2*). All tested combinations yielded between 183 and 204 Nucleocapsid N1 fragment copies per 5μL RNA extract. To characterize the extraction efficiency of the automated process, the Accuplex viral particles were directly added to 96-well PCR plates, and subjected to thermal/enzymatic treatments before performing RT-qPCR. This direct PCR method was considered as 100% (average 327 copies) and compared with other methods employed during this study. The comparative extraction efficiencies of the automated system with H_2_O, EtOH, and DRS were 61.0%, 61.5%, and 55.9%, respectively, while the manual method with DRS was 62.2%. All the extraction procedures exhibited high variabilities; however, the automated system demonstrated a lower coefficient of variation (4.0-7.7%) compared to manual kit extraction (8.4%).

**Figure 2:**
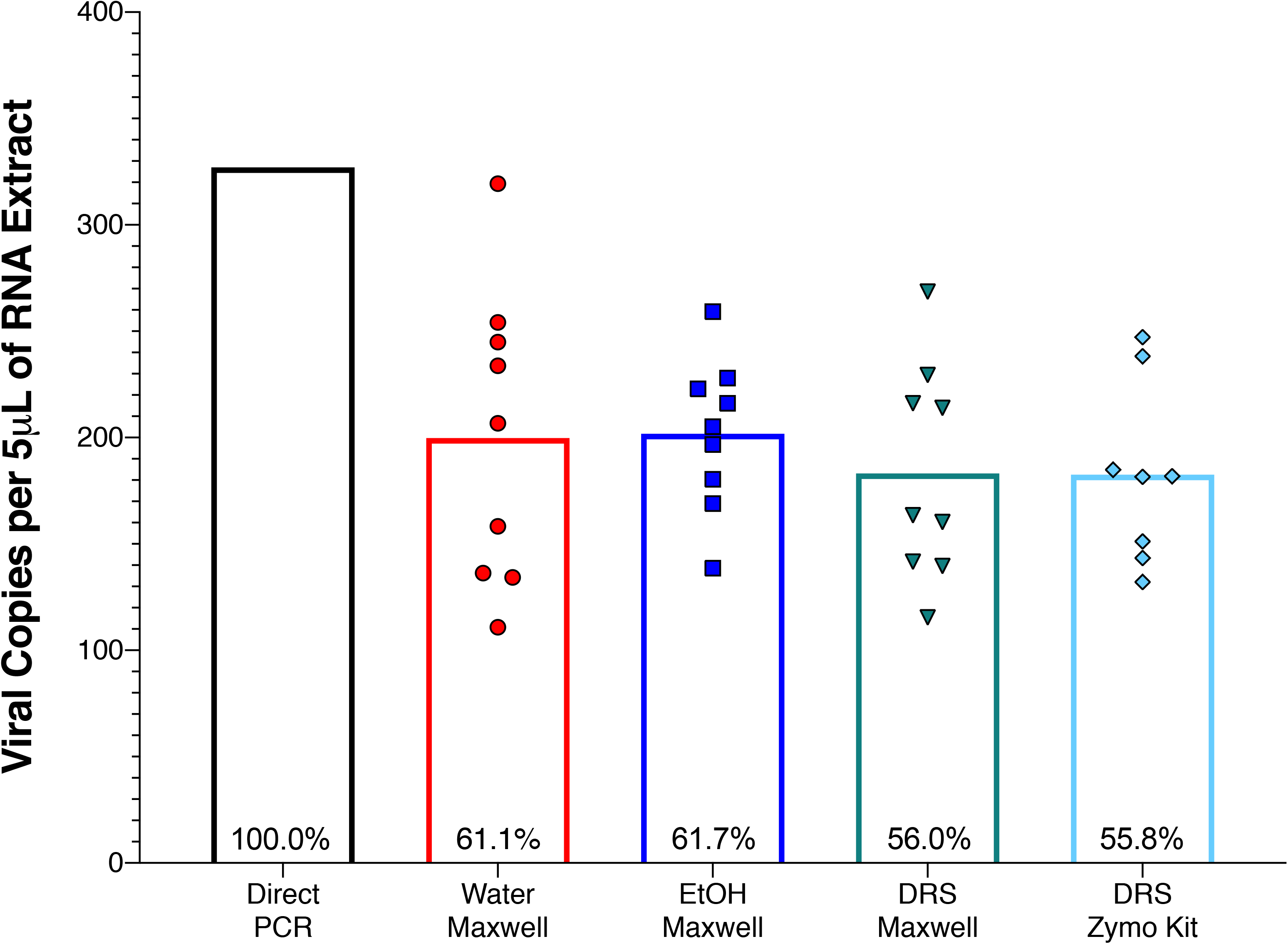
Extraction kit efficiency. RNA extraction from AccuPlex viral particles was examined using Direct PCR (black column) and compared to four different combinations of storage liquids and extraction kits including Maxwell RSC Viral extraction kit with water (●), ethanol (EtOH; ■), and DRS (▼), as well as Zymo Quick-DNA/RNA Viral Kit with DRS (♦), followed by quantification using RT-qPCR assay. Values are expressed as nucleocapsid (N1) copy numbers in 5μL of RNA extract; all replicates are plotted as individual points, with presented as columns. Direct PCR was treated as 100% to calculate the extraction efficeny of the other extraction methods (recorded with in the columns). Significant differences were determined by Welch’s t test, significance (p<0.05) denoted by ‘*’.

### E2E Assay

Zeptometrix NATtrol, an inactivated SARS-CoV-2 positive control, was used in these studies since the AccuPlex stock contains high concentrations of glycerol, making it challenging to dry onto material surfaces. For this study, 5,000 copies of NATtrol viral particles per 25 cm^2^ were spotted on bare stainless steel (BSS), painted stainless steel (PSS), polyethylene terephthalate modified with glycol (PETG), and fiberglass-reinforced plastic (FRP) materials. After desiccation of the viral control on the surface, the viral droplets left a visible plaque on all surfaces (smaller dots within the swabbed area), *Figure 3A*). Sample collection with the swab showed noticeable differences in the amount of the plaque that was dissociated during swabbing. The visible marks associated with BSS, PSS, and FRP materials remained mostly intact after swabbing; however, roughly half of the PETG plaques broke apart during swabbing (*Figure 3A*). Such plaque breakup possibly allowed for collecting larger pieces of the viral plaques on the PETG. Materials of 25 cm^2^ (coupons) were swabbed 18 hours after inoculation (Day 1) and re-swabbed (with a fresh swab) after incubating at room temperature for additional 24 hours (Day 2). On Day 8, after inoculation, the coupons were wiped down with 0.6% bleach (sodium hypochlorite) and then re-swabbed with a fresh swab for the third time.

**Figure 3:**
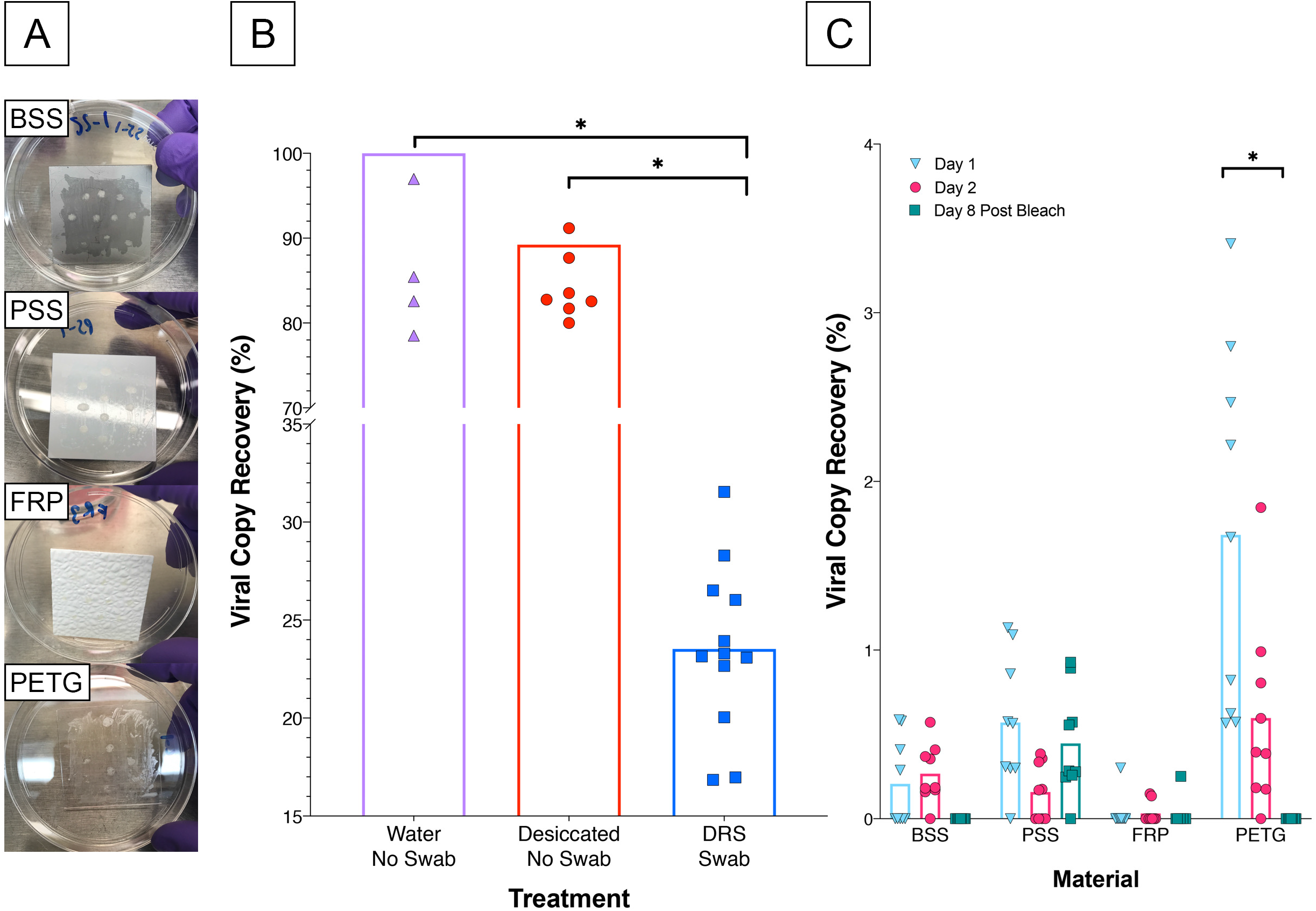
Viral particle recovery from built-environment surface materials. (A) Image of the inoculated plates (BSS, PSS, FRP, and PETG) where inactivated viral particles (10 μL of NATrol) were aliquoted 10 times on to four separate materials in triplicate. (B) Viral particles were either kept overnight at room temperature as liquid (Eppendorf tube was closed; Water No Swab, ▲) or desiccated over night at room temperature in tubes (No Swab Desiccated ●) which were then sacrificed to extract RNA directly without removing from the surface. In addition, aliquots of viral particles that were not desiccated but inoculated in DRS and swab materials were also processed (DRS Swab, ■). (C) Viral particles were collected from the seeded surfaces with Isohelix swabs and DRS, extracted on the Maxwell RSC, and quantified using RT-qPCR assay. Viral RNA copy number for each condition was divided by an extraction control to calculate percent recovery for Day 1 (▼), Day 2 (●), and Day 8 post bleach (■). Statistical significance was determined by Welch’s t test with significance (p<0.05) denoted by ‘*’.

Initially, when viral particles were desiccated in an Eppendorf tube and RNA extracted directly, a 11% loss of RNA due to desiccation was documented when compared to the solution that was not dried. The average percent recovery of the viral particles directly inoculated onto the swabs in DRS was ~23% (*Figure 3B*). However, when desiccated on materials, the highest percent of RNA recovery after Day 1 was observed for PETG material (1.68%), followed by PSS (0.57%) and BSS (0.21%) (*Figure 3C*). The lowest observed recovery was from FRP at 0.03%. (*Figure 3C*). On Day 2, viral recovery decreased on PETG (0.6%) and PSS coupons (0.16%); however, no decrease was noted on Day 2 for the BSS and FRP materials (*Figure 3C*). After treatment with 0.6% bleach, the recovery from BSS and PETG materials decreased to below detection limit (BDL), while the bleach was not effective in the removal of RNA from PSS as traces of RNA could still be detected (0.45% recovery), while only 0.03% recovery was observed on FRP. This might be because the RT-qPCR assay could detect very short fragments of RNA (~70 bp) and hence likely amplified degraded nucleic acids. This test revealed that viral persistence on surfaces varies, and in some cases (such as on PSS) viral RNA can be recovered after cleaning with bleach.

### Comparison of RT-LAMP and RT-qPCR Assays

Reverse-transcription loop-mediated isothermal amplification (RT-LAMP) is a single step colorimetric presence-absence assay that can be used as a narrow range semi-quantitative assay to determine the presence of SARS-CoV-2. When used as described by the manufacturer, the colorimetric RT-LAMP assay generates a yellow color for a positive result or remains unchanged (pink) for a negative result. Samples with borderline results have a gradient color change between yellow and pink. All samples can be further analyzed to obtain narrow range semi-quantitative results by measuring the resulting DNA product with the Qubit DNA broad range quantification kit. Since the dynamic range between positive and negative is narrow, negative reactions (pink) have final DNA concentrations of <50 ng/μl post amplification, and full-color positives have a post amplification of ~550 ng/μl in a 25ul reaction. Samples that are borderline will have DNA yields between 50 and 550. For these studies, both positive viral controls were lysed directly (*Supplemental Table 1*), and the lowest limit of detection was determined at 5 and 12.5 copies per reaction for AccuPlex, and NATrol, respectively (*Figure 4A*). Since the RT-LAMP assay is not truly quantitative, values < 150 ng μL^-1^ DNA concentration were considered negative, and values ≥ 150 ng μL^-1^ were considered positive.

**Figure 4:**
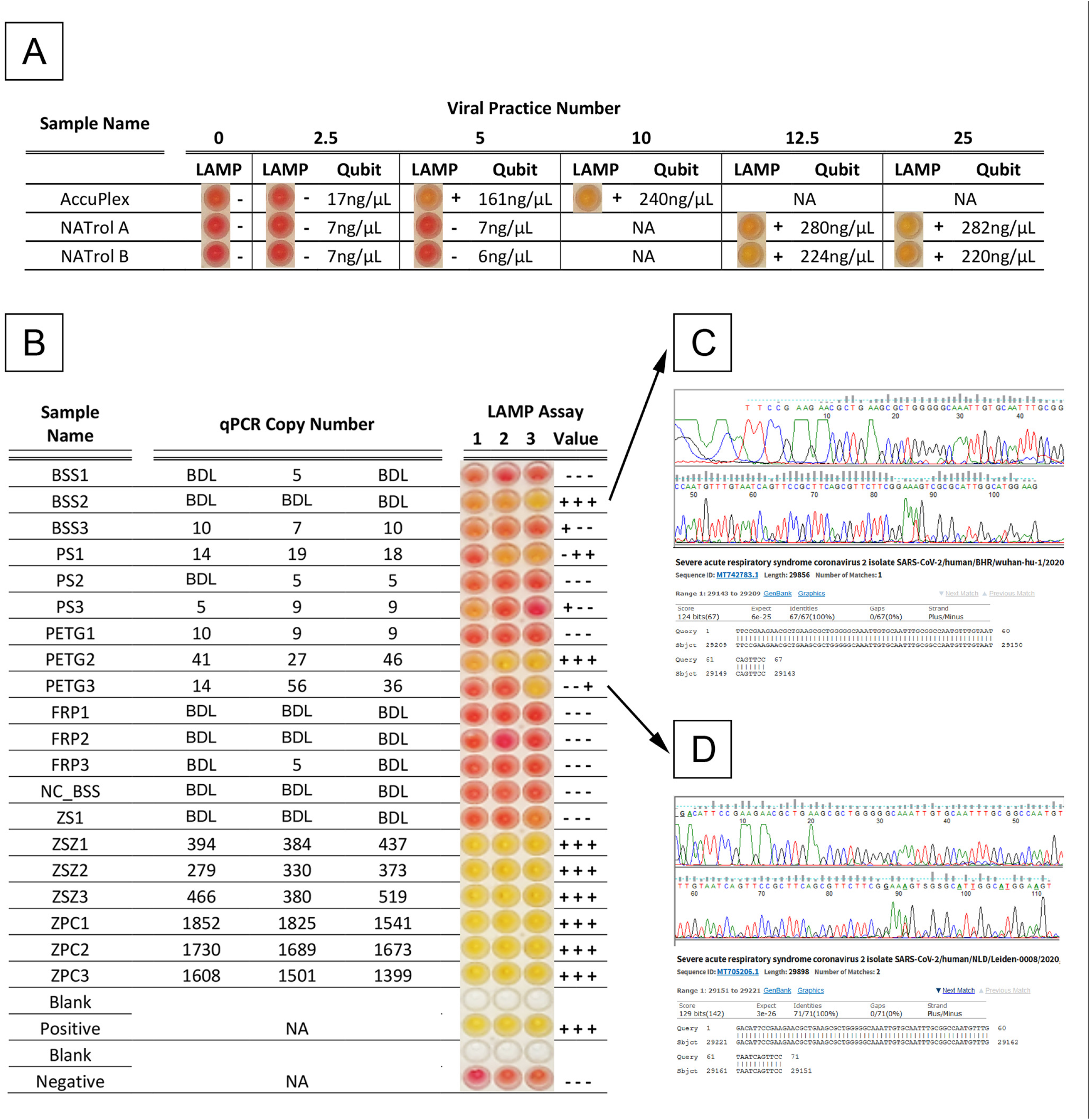
Comparison of RT-LAMP and RT-qPCR assays. (A) RT-LAMP assay limit of detection was carried for AccuPlex and NATtrol standards with both the colorimetric changes seen in the reaction (RT-LAMP Assay output) and the Qubit quantifications presented across a dilution series of viral particle number. The qualitative RT-LAMP assay output was determined based on color change from red to yellow in the presence of the target sequence, whereas RNA measurements of RT-LAMP assay reactions using Qubit give semi-quantitative values. Qubit values that were below 100 ng/μL were denoted as ‘-’ and Qubit values that were above 150 ng/μL were recorded as ‘+’. (B) Viral particles collected from built-environment surface materials (Figure 3 Day 1) were analyzed with the RT-LAMP and RT-qPCR assays. RT-LAMP Assay colorimetric output is presented alongside Qubit +/-result value, and RT-qPCR quantities. Values that were not tested were marked as not applicable (NA), and values that were undetectable were recorded as BDL.

The samples collected from the inoculated coupon (*Figure 3A*) and analyzed by RT-qPCR were further analyzed using the RT-LAMP assay (*Figure 4B*). When the RT-qPCR results were compared with the RT-LAMP assay results, >300 copies were definitively positive with RT-LAMP assay (yellow coloration). However, for samples with concentrations near the limit of detection (LOD) for RT-LAMP assay (~12.5 copies/μL), the results between RT-qPCR and RT-LAMP were less correlative. (*Figure 4B*). For samples that exhibited discrepancies between the two assays (BSS2 and PETG3), Sanger sequencing was performed and confirmed SARS-CoV-2 sequences (Figure 4C) indicating the reliability of the RT-LAMP assay.

### Materials-Associated Organics Inhibition in the RNA Recovery

Since many of the chemicals associated with cleaning, disinfection, and indigenous chemical constituents of the materials could have PCR inhibitors, we conducted several experiments to determine this potential. The precision cleaned uninoculated surface materials (25 cm^2^) described above (BSS, PSS, PETG, and FRP), were swabbed and placed in DRS media along with 5000 copies of the NATrol viral control. These samples were processed along with a positive control that included a swab in DRS media with NATtrol viral control but not exposed to any test surfaces. Results indicated that all swabs used for sample collection of surface materials demonstrated similar recovery rate as the controls not used in surface sampling. Analysis of variance (ANOVA) indicated that swabs used to sample both BSS and FRP had similar recovery rates of 25.2% and 24.3%, respectively, while PSS had 30.8% and PETG had 36.0% (*Supplemental Figure 1*). Swabs sampling from PETG had a significantly higher recovery rate than FRP (p=0.0001) and BSS (p=0.0006), while PSS was significantly higher than FRP (p=0.0152) and BSS (p=0.0439) surface types. The recovery percentages exhibited for all swabs were within a standard deviation (Average 29.5% +/−5.5% standard deviation). These recovery rates from various tested materials (24% to 36%) were similar to the positive control that were not exposed to any test surfaces (20% to 25%; *Figure 3B*), which demonstrated that the precision cleaning did not leave residual organics or debris that could inhibit the RT-qPCR assay. The difference in the recovery was attributed solely to the DRS-swab combination, since viral particles spiked in water with swab without DRS solution demonstrated an ~88% recovery.

### Influence of Environmental Debris on Viral Quantification

To determine whether the detection of SARS-CoV-2 virus would be affected by the debris associated with built environment materials (stainless-steel metal, Amerstat, plastic, copper, painted surfaces, and wood), samples were collected and efficiency of the E2E procedure was tested. The collected materials in DRS solution were inoculated with and without AccuPlex (500 copies) viral standards prior to RNA extraction and RT-qPCR assay. As expected, there was a significant inhibition in the recovery of the AccuPlex viral RNA from all surface materials swabbed, ranging from 50% recovery for stainless-steel to only 4% and 8% for Amerstat and the painted surfaces, respectively (*Figure 5*). Wood, copper, and plastic surfaces exhibited intermediate recovery of viral particles at a level of ~20%. On average, the recovery of AccuPlex viral RNA from the stainless-steel was significantly higher than all other tested surfaces (p=0.05 to 0.004), but the AccuPlex RNA recovery was more variable with a range from 15% to 100%. Barring one or two outliers, the recovery from plastics was consistent (*Figure 5*).

**Figure 5:**
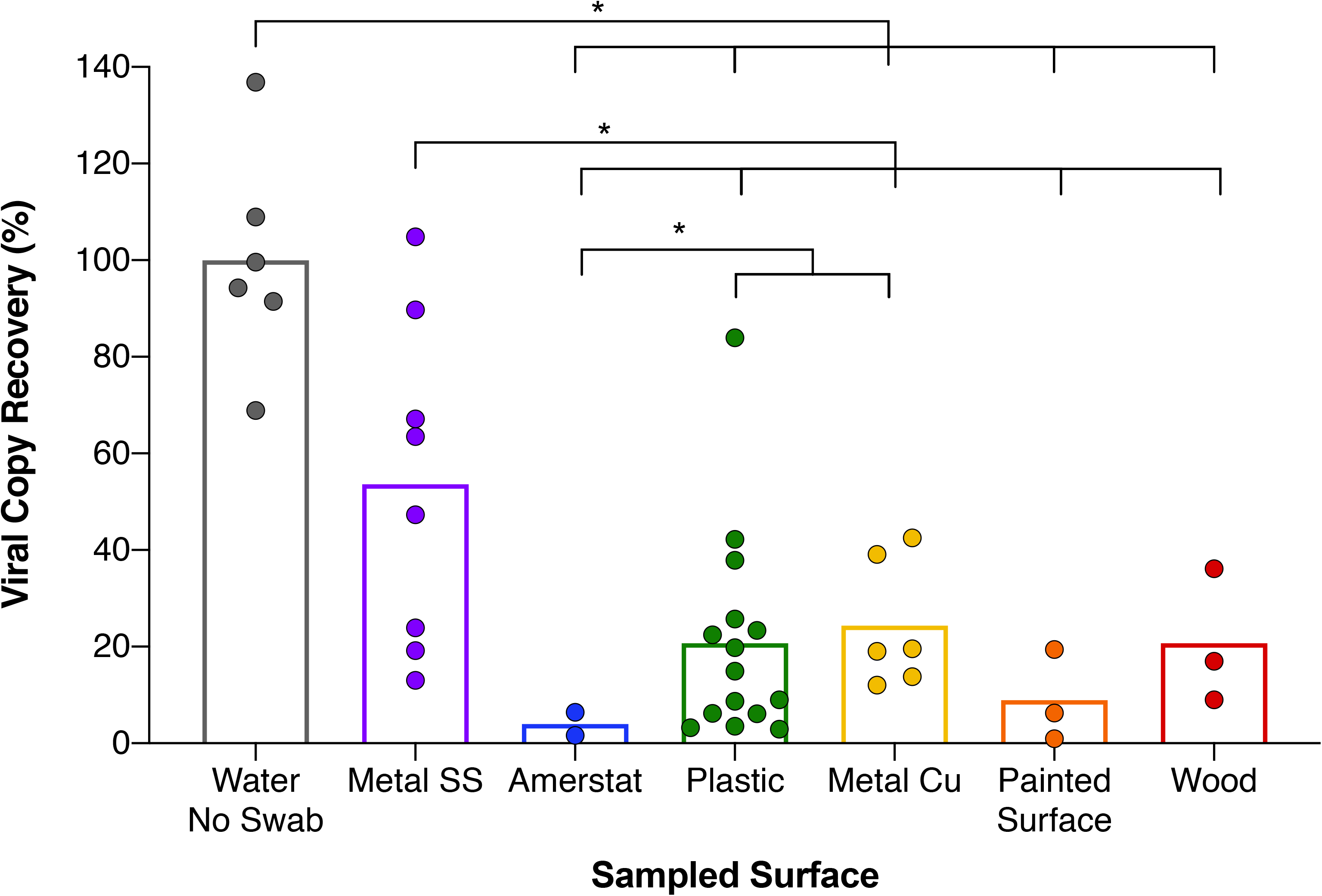
Inhibition by field collected built-environment surface samples post RNA extraction. Field swab collection of diverse built-environment surface samples had their DRS vials spiked with inactivated viral AccuPlex particles prior to RNA extraction and quantification with RT-qPCR. Differential amplification of Metal SS (●), Amerstat (●), Plastic (●), Metal Cu (●), Painted Surface (●), and Wood (●) was compared to a positive control (●) and reported as percent recovery compared to that positive control mean. Each column represents average percent recovery for respective surface type. Significance (p<0.05) denoted by ‘*’, based on Welch’s t test.

To ascertain whether the decreased AccuPlex viral RNA recovery was due to the interaction of the environmental debris with the organics in extraction reagents, the post-extract of the six materials were spiked with 500 copies of synthetic fragments obtained from Integrated DNA Technologies (IDT) prior to being subjected to RT-qPCR. In contrast to the results above, which showed large inhibition, amplification of the spiked synthetic IDT fragments after the RNA extraction was largely unaffected. The copper resulted in the lowest recovery at 77%, followed by plastic, wood, and painted surfaces (~84%), while the stainless-steel and the Amerstat exhibited 90% recovery when compared to the control (*Figure 5, Supplemental Figure 2*). These results underscore how the type of environmental surface can influence the recovery of viral molecules, while the RNA purification kit chemistry might account for a small percentage of such inhibition.

### Validation of E2E Process by an Independent Laboratory

The E2E assay was repeated using the same materials by an independent laboratory for reproducibility and verification purposes. The independent evaluation included LOD determination of RT-qPCR assay, RNA extraction efficiency of automated system/manual kits, and recovery of NATrol viral particles from various built environment material surfaces. The results of the second laboratory evaluation were comparable and or equivalent to the results presented here. A standalone report is included in Data Set-1.

### Built Environment Study Testing SARS-CoV-2 from Environmental Surfaces

The E2E protocol developed during this study could confirm viral presence from built environment surfaces only when ≥1,000 viral particles per 25 cm^2^ were present due to the losses associated with swab collection, transportation solution, RNA extraction, and material surface retention. Despite these limitations, the combination of using Isohelix swab, DRS as transportation medium, automated RNA extraction, and RT-qPCR assay was determined to be the best available E2E protocol during March 2020 to reproducibly detect and measure SARS-CoV-2 from built environment surfaces. The E2E process implemented during this study are shown in *Figure 6*. The samples collected were from seven different materials found in 10 buildings, including stainless steel, Amerstat, plastic, copper, and painted surfaces. None of the 368 samples collected tested positive for SARS-CoV-2 (i.e., RT-qPCR amplification for N1 gene was BDL) using the E2E process developed during this study. Since the detection sensitivity of the E2E process implemented was 1,000 viral particles per 25 cm^2^, the samples collected from built environmental surfaces were either devoid of the targeted virus or BDL of the E2E assay.

**Figure 6:**
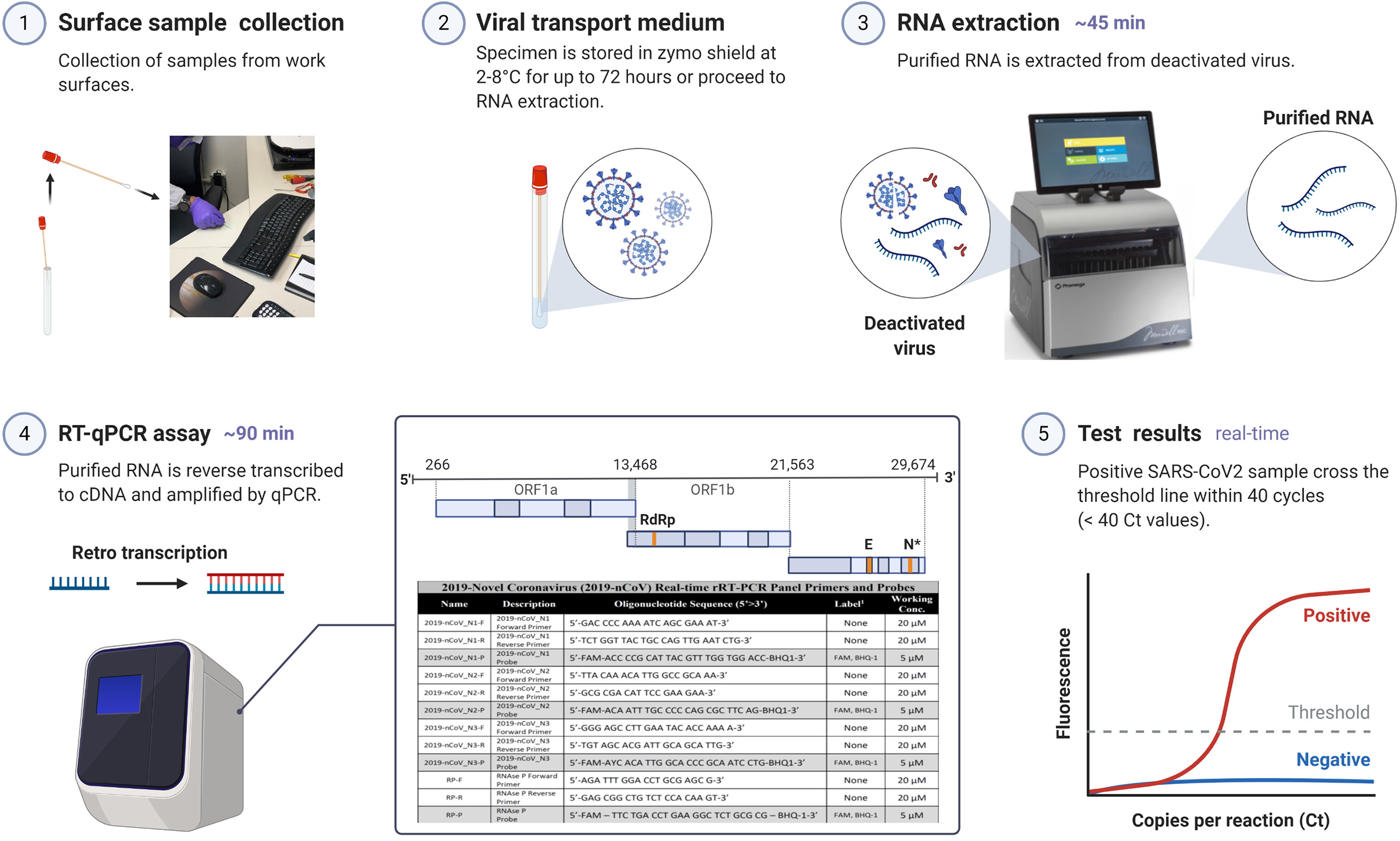
Environmental surface testing using E2E protocol. The optimized E2E protocol for detecting SARS-CoV-2 virus on surfaces is a 5 part procedure: (1) Surface sample collection; (2) Viral transport medium; (3) RNA extraction; (4) RT-qPCR assay; and (5) Test results.

## Discussion

The current clinical method for screening potential SARS-CoV-2 virus patients require an initial throat and or nasopharyngeal swab sample collection (9). Unlike clinical samples, fomites and high-touch surfaces that become contaminated with the virus display lower concentrations of the virus (10), which are often difficult to detect due to method limitations and, in some cases, inhibitory materials. For this reason, robust methods are imperative for the recovery and detection of SARS-CoV-2 from environmental surfaces. Previous studies have analyzed a variety of methods for viral recovery from surfaces (11); however, there are substantial number of variables that can impact collection, processing, and quantification of viral particles. Despite the World Health Organizations “How To” guide for SARS-CoV-2 surface sampling in hospital settings, there has not been a comprehensive study that adequately addresses all the issues associated with an E2E assay for SARS-CoV-2.

During this study, Isohelix swabs were selected over Copan swabs due to easier handling and higher sensitivity for sample collection, and this approach has been successfully used by other studies (12). Furthermore, our results demonstrated that automated RNA extraction was as efficient (13, 14) as manual kits for extracting synthetic SARS-CoV-2, which has been previously noted using phenol-chloroform (15).

At the outset of this study, in February 2020, there were several molecular methods available for assaying the virus in a given sample. Various reports demonstrated well-established techniques, such as RT-qPCR (16), RT-LAMP (1), polyA RNA-seq (17), ribo-depletion RNA-seq and MeRIP-seq (18), direct RNA sequencing (19), capture panel / amplicon (20), and digital droplet PCR (ddPCR) (21). While each of these techniques has its strengths and merits, in the context of a diverse, low-biomass sample, each have their own shortcomings, which limit their use for environmental surveillance applications. This includes elements such as detection limits, costs, inhibitor affects, input volumes, result type, or ability to validate a positive result. However, despite these shortcomings, two distinct molecular technologies (RT-qPCR and RT-LAMP assay) evolved to become part of the mainstream research toolkit for both clinical and environmental testing (add mason wired paper here). The benefits of these assays, when run in tandem, help resolve data associated with the more challenging and complex environmental sample type to accurately detect and quantify SARS-CoV-2 from various surface materials.

While comparing RT-qPCR, RT-LAMP, and ddPCR to detect SARS-CoV-2 from surface samples, ddPCR was found to be the most accurate and repeatable diagnostic tool. However, ddPCR is more expensive and requires a specialized ddPCR instrument. In contrast, the RT-LAMP assay is faster and less labor intensive than ddPCR, is relatively inexpensive, and requires minimal instrumentation to operate (e.g., a heat block, water bath or thermocycler). The results of the RT-LAMP assay are colorimetric and the low infrastructure requirements make the assay ideal for field testing as demonstrated in past studies (22, 23). Since RT-LAMP is a narrow range semi-qualitative assay, accurate quantification is best performed by RT-qPCR; the gold standard for viral RNA detection (24-26). For our studies, we selected RT-qPCR for its wide dynamic range, throughput, and sensitivity (2 copies/μL) as the primary analysis tool, with RT-LAMP as the confirmatory assay.

Once an infected person begins shedding SARS-CoV-2 viral particles, the primary route of infection is via respiration; either through droplets or aerosols that are expelled during normal speech, respiration, and especially sneezing, and unintentionally inhaled by even healthy individuals (27). Although the virus appears to be primarily transmitted through air, SARS-CoV-2 can remain viable on surfaces for up to 72 hours (6, 7). Thus, similar to other respiratory viruses (28), it is likely that a major route of SARS-CoV-2 infection comes from contact with infected surfaces followed by inadvertent touching of the face and mouth. This pattern was observed in a study in a Wenzhou, China, where numerous individuals became infected, despite not having any direct contact with known patients (29). These findings, in combination with virus longevity on surfaces, strongly suggest that transmission is not just limited to aerosols. Additionally, preliminary research suggests that the infective dose is lower for SARS-CoV-2 in comparison to other respiratory infections (30, 31). These findings highlight the importance of effective environmental surveillance, surface monitoring, and proper sanitization methods to eliminate the virus.

In this study, we analyzed several environmental surface materials that were inoculated with a known concentration of SARS-CoV-2 viral reference standard to determine the recovery efficiencies for each material. While each material has characteristics that contribute to recovery, surface roughness and hydrophobicity are important contributors. It has been reported that surface roughness is a key mitigating factor for lower recovery of biological materials (32). For the test surfaces evaluated in this study, FRP had a textured surface and resulted in a lower NATrol viral recovery. Even though the PETG surface had similar surface roughness as BSS and PSS, the surface texture of PETG might have enabled the recovery of more virions. This was evident from visual observations revealing that the dried virus inoculum was easily dissociated and resulted in higher recoveries-a likely result of both the smoothness and hydrophobicity.

The decreasing amount of recoverable NATrol viral particles over time from the surface materials are likely attributed to the combination of desiccation time (Day 1 to 8) and the use of a disinfectant. Unlike all other materials, PSS demonstrated nearly the same RNA copy numbers persisting from Day 1 desiccation until Day 8 post-bleach. This might be due to the RT-qPCR method, which was targeting only 67 to 71 bp amplicons that could still easily be detected from virion fragments degraded by the disinfectant. Despite desiccation, the chemical nature of the paint associated with PSS material might have allowed viral fragments to persist even after cleaning with bleach. Furthermore, after applying bleach, the pigmentation of the paint was altered indicating a chemical reaction had occurred, which may have enabled easier removal of viral particles from the surfaces. Similarly, recovery of viral fragments was documented on the cruise ship *Diamond* after hypochlorite disinfection of contaminated rooms (33). These surprising positive results were likely due to the detection of degraded RNA fragments still being detected in the short amplicon RT-qPCR method. Due to short fragment amplification, even significantly degraded RNA can be detected. In order to avoid these false positive results after the bleach treatments, samples should be tested using an alternative technique that targets longer RNA fragments, such as RT-LAMP.

To detect the high concentration of SARS-CoV-2 virus in clinical samples, it has been shown that direct amplification was possible with a maximum sample input of 2 μL (34), whereas for environmental samples, a RNA purification step was mandatory due to the PCR inhibitory substances (13, 35). In addition, the concentration of the target molecules during RNA extraction would allow larger volume input (10-fold more) and increased the detection limit (2 copies per μL of RNA extract). The collection of microorganisms from environmental surfaces have been documented to yield ~1 to 10% of biological materials due to issues in their removal from the surfaces as well as challenges associated with their dissociation from the swab (13, 35). During our study, the DRS chemistry in combination with environmental debris and RNA extraction has compounded losses an additional ~80%.

The E2E process implemented to survey SARS-CoV-2 virus presence for built environment surfaces (n=368 samples) exhibited no viral incidence (or <1,000 viral particles per 25 cm^2^), which might be attributed to a highly controlled practices that were strictly adhered. These practices included but were not limited to admitting limited number of employees at a given time period, training “Safe at Workplace,” enforcing social distancing, wearing masks, practicing personal hygiene, and deep-cleaning of the environmental surfaces might have limited the viral contamination in these built environment surfaces. However, high traffic areas like hospitals, restaurants, cruise ships, and subways might show a different pattern(s) of viral adherence and persistence on fomites and surfaces (33, 36).

## Conclusion

When examining all elements, the optimized E2E protocol implemented during this study indicated that only ~0.5 to 2% of the viral particles could be recovered from a variety of built environment surfaces and a minimum of 1,000 target molecules (viruses) per 25 cm^2^ were needed to positively detect the virus. During this study, it was established that 1% of NATrol viral particles were recovered due to sample collection (swabs) and transportation solution (DRS), and that the RNA extraction step accounted for a further 90% loss of target molecules. These data reflect an overall E2E process efficiency of 0.1%, meaning that at least 1,000 copies need to be present for successful and reproducible detection of the SARS-CoV-2 virus from environmental surfaces.

## Methods

### Inactivated Viral Reference Standards

Two noninfectious, replication-deficient, encapsulated SARS-CoV-2 viral reference standards were used during this study, including the SeraCare AccuPlex (Milford, MA; Cat#: 0505-0126), which contained the ORF1a, RdRp, E, and N sequences, and the ZeptoMetix NATtrol (Buffalo, NY; Cat#: NATSARS(COV2)-ERC), which contained the entire RNA sequence. The AccuPlex and NATtrol stocks were purchased at a concentration of 5 x 10^3^ and 5 x 10^4^ viral particles per mL, respectively. These concentrations were confirmed in-house using digital droplet PCR (ddPCR) to be within 1.25% accurate (*Supplemental Table 1*).

Digital droplet PCR was performed using the BioRad QX200 instrument with the IDT primer/probe set for N1 and N2 with a modified probe quencher of Iowa Black ZEN/IBFQ (Cat# 10006770) along with the BioRad One-Step RT ddPCR advanced supermix (Cat #1864021). Four methods were used for extraction of RNA from these reference materials (i.e., AccuPlex and NATrol) and consisted of the following: (a) direct lysis at 75°C for 5 min, (b) direct lysis of a 1:1 mixture of sample to nuclease-free water (15ul:15ul) to which 3ul of Proteinase K (Qiagen) and 0.8ul of RNase inhibitor (Ribolock, Thermo Scientific E00381) was added and incubated at 50°C for 10 minutes followed by freeze thaw −80°C to + 95 °C for 4 minutes, (c) utilization of viral RNA extraction kits such as the QIAamp Viral RNA Mini Kit (Qiagen, Germantown, MD; Cat #52904) and (d) the RNeasy Micro kit (Qiagen; Cat #: 74004). Volumes of 1, 2, 3, 5, and 7 μL of the Accuplex and NATrol viral standards were analyzed using methods 1 and 2 for ddPCR to determine exact copy number.

### Swab and Viral Transport Medium Selection

Two protocols, involving sample collection and transport medium, were tested during this study. The first protocol in *Supplemental Figure 3A* shows the procedure used for the Metagenomics and Metadesign of Subways and Urban Biomes (MetaSUB) and heritage NASA environmental sampling (12, 37, 38). The Copan Liquid Amies Elution Swab (ESwab, Copan Diagnostics, Cat.:480C) were used for environmental sampling. Sampled Copan swabs were stored on dry ice and transferred to the lab for further processing. Once in the lab, 300 μL of lysis buffer and 30 μL Proteinase K (Promega, Madison, WI) were added, and the swab was cut using sterile scissors to release the swab into the tube and mixed thoroughly using a vortex. The materials released from the swab were extracted using the Maxwell Viral Total Nucleic Acid Purification Kit (AS1330; Promega) or Zymo Quick-DNA/RNA viral kit (Cat# D7020). The protocol shown in *Supplemental Figure 3B* represents the reference protocol procedures used for a similar study design by the 2017-2019 MetaSUB research consortium (12). This process used the Isohelix MS-02 swab (Mini-Swab, Isohelix Cat.:MS-02) with 400 μL of DRS (R1100-250) preservative. Sampled Isohelix swabs were broken off into the sample tube and transferred to the lab at room temperature. Once in the lab, samples were extracted for nucleic acids via the Promega Maxwell RSC 16. Among the swabs tested, Isohelix swabs demonstrated higher a recovery of microorganisms compared to the Copan swabs (39). Since no published reports were available on the efficiency of swabs specific for virus collection from environmental surfaces, data from the MetaSUB consortium (39) were adapted for this study. The DRS medium used throughout this study (DNA/RNA Shield -Zymo Corp) contains proprietary chemicals that inactivate the live virus and preserve RNA at a biosafety level 2 status.

### Efficiency of Various Protocols in Extracting Viral RNA

The standard methodology for viral RNA extraction in this study involved using the surface samples collected in DRS (~200 μL), and processing them using the automated Maxwell RSC extraction platform (Promega Corp., Madison, WI) following the manufacturer’s instructions for Maxwell RSC Viral Total Nucleic Acid Purification Kit (Promega). In brief, the collected swabs were vortexed for 2 min and treated with the lysis solution provided by the manufacturer (220 μL of the lysis buffer per 100 μL of sample and 200 μL of the DRS solution). This extraction tube was incubated at room temp for 10 minutes and 56°C for additional 10 minutes. Samples were transferred to Maxwell cartridges for extraction using the Viral Total Nucleic Acid program of the instrument. Purified RNA was eluted into a 60 μL of UltraPure molecular grade water and divided into two aliquots. Samples were stored at −80°C with one aliquot used for downstream RT-qPCR analysis while the other aliquot was archived for later use.

In order to compare the efficiency of the extraction protocols and the effects of the DNA/RNA Shield on RNA amplification, four sets of extraction fluids were prepared in triplicate. Set one was prepared with 100 μL of AccuPlex in 100 μL of UltraPure water; set two was prepared with 100 μL of AccuPlex in 100 μL 95% EtOH; and sets three and four were prepared with 100 μL of AccuPlex in DRS. Sets one, two, and three were all processed on the Maxwell RSC as described above. Set four was processed using the Quick-DNA/RNA Viral Kit (Zymo Research, Irvine, CA) following the manufacturer’s protocol. Purified RNA was eluted in 60 μL of UltraPure water.

### Synthetic RNA and Limit of Detection for RT-qPCR

Two synthetic nucleic acid reference samples were used to generate standard curves for the RT-qPCR reactions: (i) 2019-nCoV N Positive DNA Control (10006625) from Integrated DNA Technologies (IDT) (40) and (ii) SARS-CoV-2 RNA Control 2 (MN908947.3) from Twist Biosciences (San Francisco, CA). The IDT standard consisted of control plasmids containing the complete nucleocapsid gene from SARS-CoV-2, while the Twist standard consisted of six synthetic 5kb ssRNA section of the viral genome. Both IDT and Twist contain the nucleocapsid gene and can be amplified by either N1 or N2 primer sets, producing amplified products that have lengths of 72 bp or 67 bp, respectively. Comparison of N1 and N2 primers using the IDT and Twist BioSciences synthetic standards showed that all combinations of the primers and standards had highly reproducible amplification quantities across log dilutions, with N1 demonstrating a slightly higher efficiency amplification curve. (*Supplemental Figures 4A—E*). Hence, only the N1 primer set was used for developing the E2E protocol. Samples that resulted in a N1 positive results were further confirmed with the N2 primer set. A significantly higher viral copy number was detected using the IDT reference material (1.28-fold) in comparison to Twist (P < 0.05) when assessed with RNA extracted from AccuPlex as a benchmark control (*Supplemental Figure 4E*).

To determine the limits of detection of the RT-qPCR assay, a two-fold dilution series from 0 to 200 viral RNA copies per reaction volume (5μL; 12 replicates), were conducted and indicated a LOD of 10 viral RNA copies per 5 μL reaction volume (2 copies/μl; *Supplemental Figure 5A*).

Among the 12 replicates that theoretically contained one viral RNA copy (5 copies/5μl), five did not reach the cycle threshold (Ct) and were thus considered as BDL. All no template controls (NTC) were negative. As expected, the standard deviation of Ct values increased as the molecule concentration decreased (<10 copies) (*Supplemental Figure 5B*).

### Optimization of RT-qPCR Assay

qPCR was carried out with the extracted viral RNA from the sample using the Luna Universal Probe One-Step RT-qPCR Kit (#E3006, New England BioLabs [NEB], USA) as per the manufacturer’s protocol for Applied Biosystems real-time instruments. N1 and N2 IDT primers (2019-CoV CDC EUA Kit, Integrated DNA technologies) designed for CDC SARS-CoV-2 qPCR probe assays were used for all reaction setups. The kit consists of all published SARS-CoV-2 assays in the CDC’s recommended working concentration. The final 20 μL reaction mix also included Antarctic Thermolabile UDG (Uracil-DNA Glycosylase) to prevent sample cross contamination. The IDT SARS-CoV-2 Plasmid DNA Control was used to generate a log_10_ standard curve from 1 to 10^5^ copies in triplicate. The AccuPlex SARS-CoV-2 reference material was used as an extraction control and treated as an “unknown” sample for each analysis. A QuantStudio 6 Flex Real-Time PCR Detection System was used for all RT-qPCR runs.

Cycling conditions were: reverse transcription, 55°C (10 min, 1X); initial denaturation, 95°C (1 min, 1X); and 40 cycles of 95°C (15 sec), 60°C (60 sec) plus plate read. The N1 gene was used to determine the number of viral particles in a sample. NTCs, a reaction mixture with molecular-grade water substituted for the sample, were run on each RT-qPCR plate to serve as negative controls. Standard curve and quantification were carried out using the Design and Analysis Software Version 2.4.1, for QuantStudio 6/7 Pro systems.

There are some unresolved issues with the RT-qPCR, which repeatedly detected higher amounts of AccuPlex and NATrol in standard controls than the quantity that was being measured by dd-qPCR. Supplemental Figure 6A-B shows the number of copies per mL of AccuPlex and NATrol that were calculated based on RT-qPCR runs. The red lines demarcate the reported concentration of viral particles for AccuPlex and NATrol. Supplemental Figure 6C shows that there was a 2.69-fold higher concentration of viral particles for AccuPlex and 3.5-fold higher for NATrol.

### RT-LAMP Assay

A 5 μL aliquot of each sample was analyzed in triplicate using the RT-LAMP assay with the WarmStart RT-LAMP reagent (M1800S NEB Inc Ipswich MA) and the N2/E primer mix against the nucleocapsid envelop protein gene. A custom primer mix for the final primer mix included 40 mM guanidine hydrochloride which increased the sensitivity as previously described (41). All samples were incubated at 65°C for 42 min and photographed. Sample were quantified using spectrofluorimetry with the (Qubit. Broad Range DNA kit; ThermoFisher Waltham, MA) Titrations were performed on both AccuPlex and NATtrol viral particles for estimating copy numbers. Direct RNA extraction was performed by mixing viral reference particles at a ratio of 1:1 (15 μL to 15 μL to water) and adding 1 μL of an RNase inhibitor (RiboLock-ThermoFisher EO0381) and 3 μL of Proteinase K (Qiagen Germantown Maryland), with incubation at 50°C for 10 min followed by immediately freezing at −80°C. After freezing, the controls were immediately incubated at 95°C for 4 min, followed by duplicate titrations into the RT-LAMP reaction master mix and incubation at 65°C for 42 min.

### Surface Materials Tested and Coupon Fabrication

Four of the most common high-touch surface materials were used in this study including bare 302 stainless steel (BSS), painted 302 stainless steel (PSS; white acrylic paint 168130-Rust-Oleum, Vernon Hills, IL), polyethylene terephthalate modified with glycol (PETG), and fiberglass-reinforced plastic (FRP). All materials were smooth on a macroscale, except for FRP, which exhibited an irregular, textured surface. These materials were fabricated as test “coupons” of 25 cm^2^ square at Jet Propulsion Laboratory (JPL) and sand tumbled to deburr. Throughout, coupons were handled carefully as to limit surface damage and scratching.

### Precision Cleaning of the Test Coupons

Unless otherwise indicated, all precision cleaning was performed in a Class 100 biohazard hood or a Class 100 laminar flow bench. Care was taken in handling, and high-grade chemicals were used to minimize contamination. Coupons were precision cleaned based on each individual material’s best practices, as outlined in JPL’s standard protocols (42). In short, BSS was cleaned per JPL D-51981 type IV (subsequent baths of solvent, detergent AquaVantage 815 GD; Brulin Holding Company, Indianapolis, IN), alkaline, and final passivation). The passivation consisted of a 30-min exposure to 5 M nitric acid at 24°C. Due to the paint’s associated chemical attributes and susceptibility to solvents, PSS was rinsed with deionized water. The FRP wood laminate and PETG were both cleaned per JPL D-51981 type V method C (solvent bath followed by deionized water rinse). After cleaning, the product cleanliness level was tested to the level 100 (which means particles of <0.5 μm not exceed 100 particle counts). The cleaned test coupons were individually sealed in an antistatic Amerstat bag, until use.

### Inoculation of Surface Materials (Test Coupons)

Precision-cleaned coupons were opened aseptically in a biosafety cabinet and placed into individual, sterile Petri dishes. Aliquots of 10 μL of the NATrol control were spotted (n=10) onto each test coupon in evenly spaced rows of 3, 4, and 3 spots and covered with a lid. Triplicate coupons of each of the following material types were prepared, including several controls: (i) a BSS coupon remained uninoculated (NC BSS) and were processed alongside as a negative control; (ii) a swab negative control in DRS; (iii) a swab with 5,000 copies of NATtrol in DRS; and (iv) 5,000 copies of NATtrol control extracted directly from Maxwell. All test coupons were loaded into a modified GasPak System, Anaerobic Jar 150 LG (Cat#: 260607; Becton Dickinson, Franklin Lakes, NJ) with the palladium catalyst removed and a valve port drilled into the top of the lid; no desiccation beads or reactants were added. The lid port was hooked up to a vacuum line to provide negative pressure on the jar. Coupons were dried at room temperature for 18 hours, sampled, and immediately extracted for RNA (Day 1; initial collection). After initial swabbing, coupons were stored at room temperature for 24 hours at standard pressure, and swabbed again with a fresh swab, followed by viral RNA extraction (Day 2; secondary collection). Coupons were subsequently stored at room temperature and standard pressure for another 5 days, after which coupons were treated with 10% bleach (0.6% v/v sodium hypochlorite) using Kimwipes (Kimberly-Clark, Irving, TX) using a wiping pattern vertically, horizontally, and diagonally. Coupons were allowed to dry for 30 min, swabbed, and extracted for RNA (Day 8; collection after bleach treatment).

### Sample Collection from Coupons or Built Environmental Surfaces

Test coupon were sampled over a 25 cm^2^ area using Isohelix MS-02 buccal swabs (Cell Projects, Kent, UK). Prefilled 2 mL tubes containing 200 μL of DRS and labeled with a unique barcode (Cat. No.: R1100-96-1) were used for each sample. The Isohelix swab was dipped into DRS solution for 15 s prior to sampling to ensure the swab was sufficiently moistened. The moistened swab was then held against the sample surface at a 45-degree angle and dragged in a raster pattern across the 25 cm^2^ area. To ensure good coverage over the sample area, the raster pattern was repeated three times in different directions (horizontal, vertical, and diagonal), rotating the swab head 180 degrees between the horizontal and vertical passes. The swab was held to the surface perpendicular to the direction of travel to ensure that the maximal surface area was covered by the swab during each pass in the raster pattern. After sample collection, each swab head was transferred into the same barcoded tube used to pre-moisten the swab by aseptically breaking and twisting the head off into the tube. Sample specific metadata (e.g., surface type and finish) were recorded for each barcoded tube. Environmental sampling of built environment surfaces was conducted in an identical manner. Environmental samples and field control samples were collected in a similar manner, but instead of dragging the moistened swab across a surface, the moistened swab for the field control was waved in the air for 2 min prior to breaking the head off into a barcoded tube. After collection of all samples, DRS collection tubes were stored at room temperature for up to three hours before RNA extraction.

### Environmental Debris in the Recovery of Viral Particle/RNA

Various materials, including metal, Amerstat, plastic, wood, copper plate, and painted surface, were tested to assess whether the debris associated with the environmental surface affected the recovery and detection of the viral RNA. Each surface was sampled with two swabs, which were preserved in DRS. The DRS from both collection tubes corresponding to one type of the environmental surface was pooled together, mixed, and divided into two 200 μL aliquots.

One aliquot was inoculated with 100 μL of AccuPlex control and the other with 100 μL of UltraPure water and extracted using the Maxwell RSC using the protocol for the RT-qPCR analysis. The percent recovery for each tested material was determined when compared to the control containing 100 μL of AccuPlex SARS-CoV-2 in 200 μL of UltraPure water. Three technical RT-qPCR replicates of biological samples were used for the analysis.

### Inhibition of Maxwell Extraction Chemistry in the Recovery of Viral Particle/RNA

Test surfaces were evaluated to determine if any RNA extraction inhibition was observed for the AccuPlex viral particles using the Maxwell RSC system. In order to determine the potential effect of the Maxwell RSC kit, 500 copies of the IDT synthetic fragments (2019-CoV N Positive Control, Integrated DNA Technologies, Inc., San Jose, CA) were added to each test reaction and 5 μL of each extracted sample. 20 μL of the Luna master mix (Cat#: E3006; NEB, Ispsich MA) was added, the plate sealed and analyzed using the QuantStudio 6F instrument. The percent of the inhibition for each tested material was determined by comparing to the control containing 500 copies. Three technical RT-qPCR replicates of biological samples were used for the analysis.

### Materials Associated Debris and Chemistry Inhibition

In order to determine whether debris or chemistry associated with the built environment surface materials contain PCR inhibitors, uninoculated test coupons were swabbed following the standard swabbing procedure outlined above. The swabs were transferred to a tube containing DRS and 100 μL of the NATtrol viral particles. Appropriate negative and positive controls were also included. Sample were then extracted for RNA using the Maxwell RSC and were analyzed via RT-qPCR. Inhibition was determined by comparison of the extraction ratios between positive control and the sample reaction mixtures.

### Development of SWAB Metadata Generation

For the field data collection and associated metadata characteristics of samples, JPL Information and Technology Solutions Directorate created a custom mobile application (Safe Workspace Analysis Barcode Scanner, or SWABS) that uses an iPhone to capture tracking metadata at each stage of the sample collection and analysis process. The vendor-provided barcode of each sample container was chosen as the unique identifier of each sample tracked through each phase of analysis. In the first phase (sampling), the container barcode was scanned and relevant metadata such as the name of the collection personnel, location, surface material properties, time of collection, material lot numbers, and optional description details were recorded for each sample. To accelerate data entry and eliminate human errors, barcode scanning with the iPhone camera was used to exactly identify each sample, and data in common were retained and automatically reused. In the second phase (RNA extraction), the sample was scanned once again, and details of the Maxwell machine identifier, extraction tip size, and lot numbers were added. For the third phase (archival), analysts added details of the cryo box location, extraction tip size, and lot numbers for each sample. At the final qPCR stage, the record was completed with the qPCR machine identifier, extraction tip size, and lot numbers, as well as the Ct score and copy number determined by the analysis. At each stage of the analysis procedure, the operations procedure version number was also recorded to track which documented procedure was followed at the time when each sample was processed. All of these sample processing data were gathered and stored in a centralized database at JPL. Leveraging this database, the SWABS web application makes these data accessible for viewing, searching, or editing, as well as providing reporting capabilities to communicate and summarize any of the data on demand. Although the SWABS application streamlines and improves the accuracy of the processing metadata recording process as a whole, it is most advantageous during the initial sample collection phase owing to its mobile platform that allows its users to move around freely within the workspace environment while minimally encumbered by support equipment.

### Statistical Analyses

All statistical analyses were performed using GraphPad Prism Version 8.2.0 (GraphPad Software, San Diego, California USA). Specifically, Welch’s t-test and a two-way ANOVA followed by a post-hoc sample correction were computed. Outliers were screened using the rOut method from the robustX R package (https://CRAN.R-project.org/package=robustX).

## Data Availability

There are no sequence data generated.

## Acknowledgments

Part of the research described in this publication was carried out at the Jet Propulsion Laboratory, California Institute of Technology, under a contract with NASA. Researchers associated with Biotechnology and Planetary Protection Group at JPL are acknowledged for their facility support. The authors would like to thank Garry Burdick and Subbarao Surampudi (initiating the concept of COVID-19 surface testing), Soren Madsen (day to day management of the work flow), Roger Gibbs, Leon Alkalai, Timothy O’Donnell, and the JPL Management Council (financial support and directions), Mimi Ton (IRB clearance), Anton Ovcharenko (safety protocol development), Kerry Wisden, Marisa Gamboa, and Bill Kert (procuring chemicals and materials), Oscar Rendon Perez (coupon material preparation and precision cleaning). We would also like to thank Brent Mcwatters, Mark Powell, and the members of the JPL Information and Technology Solutions Directorate that rapidly created an iPhone app that was used for metadata collection during this project. We are indebted to the personnel involved in JPL shipping / receiving for their help during this pandemic and the facility managers associated with the surface collection locations. Additionally, Mikael Kubista from the TATAA Biocenter for helping to develop our RT-qPCR limit of detection protocols, thanks to MetaSUB consortium members (especially Benjamin Young) for generating the sample collection SOP, and Julie Dragon for RT-LAMP assay SOP and continued support. Christopher Fleming and Frank Tansley are acknowledged for their timely support of procuring critical equipment and consumables. David Lee from JPL is thanked for critically reviewing the manuscript. © 2020 California Institute of Technology. Government sponsorship acknowledged.

## Ethics approval and consent to participate

Not applicable.

## Availability of data and materials

Not Applicable.

## Disclaimer

This manuscript was prepared as an account of work sponsored by NASA, an agency of the US Government. The US Government, NASA, California Institute of Technology, Jet Propulsion Laboratory, and their employees make no warranty, expressed or implied, or assume any liability or responsibility for the accuracy, completeness, or usefulness of information, apparatus, product, or process disclosed in this manuscript, or represents that its use would not infringe upon privately held rights. The use of, and references to any commercial product, process, or service does not necessarily constitute or imply endorsement, recommendation, or favoring by the US Government, NASA, California Institute of Technology, or Jet Propulsion Laboratory. Views and opinions presented herein by the authors of this manuscript do not necessarily reflect those of the US Government, NASA, California Institute of Technology, or Jet Propulsion Laboratory, and shall not be used for advertisements or product endorsements.

Work for this study way also completed by Biotia, Inc. Biotia and its employees make no warranty, expressed or implied, or assume any liability or responsibility for the accuracy, completeness, or usefulness of information, apparatus, product, or process disclosed in this manuscript, or represents that its use would not infringe upon privately held rights.

## Authors’ contributions

KV coordinated with all authors in designing the concept, executed the study, implemented the project, involved in the data analyses, and wrote the manuscript. CWP was involved in establishing the RT-qPCR assay with NKS, and was instrumental in executing the E2E process, and wrote the manuscript along with KV. NKS carried out the QA/QC of RT-qPCR assay and analyzed the data. ST helped design and setup the experimental methods, provided training, conducted RT-LAMP, qPCR, ddPCR, interpretation of data, and assisted in writing and editing the manuscript. PL carried out lab work pertaining to protocol optimization of RT-LAMP, qPCR, and ddPCR assays as well as Sanger sequencing. NKS, JMW, CWP, RH, and KC carried out environmental sampling from various built environment surfaces. JMW managed metadata collection and curation, helped analyze data, and was crucial in surface sampling as well as drafting portions of the manuscript. AB performed RNA extraction of the collected samples, contributed to data analysis, interpretation, and drafted parts of the manuscript. CU helped analyze the data, write the paper, and assisted in the study design. RH helped with sample collection and contributed in writing about the swab and DNA extraction methodology process selection for the paper. KC planned and coordinated built-environment sampling and supported data collection during sampling. AS and PV carried out RT-qPCR assays and contributed to the LOD determination. BC oversaw the project and provided critical input for the QC/QA analyses of RT-qPCR data, and edited the manuscript. NO coordinated work at Biotia, where a second laboratory performed verification out, helped design of the additional procedures used. MCR performed experiments, interpret analyzed results, and contributed to writing validation portion of the Data Set-1. MCR helped design additional experiments, performed experiments, analyzed results and contributed to writing the validation portion of the Data Set-1. DB conducted experiments pertaining to the selection of swabs and DRS and wrote these portions in the manuscript and reviewed the manuscript. CM coordinated with KV and helped to design the concept, provided insights about the MetaSUB protocol to get implemented.

## Consent for publication

All authors that participated in this study have reviewed the results, read the final manuscript, and given their consent for publication.

**Tables**

NA

## Supplemental Legends

Supp. Fig 1: <u>RT-qPCR Inhibition by Built-Environment Surface Materials</u> Uninoculated Built-Environment coupons were swabbed with Isohelix swabs and DRS, and then spiked with NATrol. RT-qPCR was performed after RNA extraction on the four sampled surface materials BSS (●), PSS (■), FRP (▼), PETG (♦), and water no swab (▲). Data is presented as percent RNA recovery for each material type as compared to a control inhibitor free NATtrol RNA extraction. All replicates are plotted and columns represent mean percent recovery. Significant differences were determined between materials using Welch’s t test, significance indicated by ‘*’.

Supp. Fig 2: <u>RT-qPCR Inhibition by Field Collected Built-Environment Surface Samples Post RNA Extraction</u> RNA was extracted from the same diverse set of Built-Environment field samples as in Fig. 6. SARS-CoV-2 cDNA was added to each sample prior to RT-qPCR to determine the inhibitor carry-through impacts on amplification of Metal SS (●), Amerstat (●), Plastic (●), Metal Cu (●), Painted Surface (●), and Wood (●) and positive control (●). Data is presented as percent recovery compared to the positive control mean, while means for each surface type are presented as columns. Welch’s t test determined significant differences between samples (‘*’).

Supp. Fig 3: <u>Collection Tool Selection and Use of Transfer Media</u> Comparison of the MetaSUB sampling SOP of 2016 and 2017-19. A) SOP for MetaSUB sampling day 2016. Samples had to be transported on dry ice and kept cold at all times. Copan swab heads have been treated with lysis buffer, cut into a 1.5ml tube containing a filter, centrifuged to separate the lysate from the foamy swab, and combined with its sample associated transport media. Those swab-specific treatments add approximately 1.5h per 96 samples to the extraction process. DNA extraction was performed using Promega Maxwell. B) SOP for MetaSUB sampling days 2017-19. Samples have been collected in DNA/RNA preservative (Zymo Shield) at room temperature. Isohelix swabs release species after lysis and vortex due to their hard surface. Samples collected in early 2017 have been extracted using the Promega Maxwell system while DNA extraction for samples received after mid-2017 has been outsourced to Zymo Research.

Supp. Fig 4: <u>Comparison of Primer Sets and Viral Standards</u> CDC/NIH recommended SARS-CoV-2 nucleocapsid N1 (red) and N2 (blue) primer sets were tested in combination with both SARS-CoV-2 standards, IDT cDNA (♦) and TWIST RNA (●), over a log dilution from 10,000 copies to 1 copy (n=5) (A-D). The N1 primer set (A, C) had lower r^2^ values than N2 primer set (B, D); however, the IDT standard with both N1 and N2 primer sets (A, B) had higher r^2^ values than the TWIST standard with N1 and N2 primer sets (C, D). The N1 primer set was used with IDT (●) and TWIST (■) to produce RT-qPCR standard curves that were intern used to quantitate five AccuPlex RNA extraction controls (S1-S5) amplified from 5 μL of extracted RNA (B). The number of N1 RNA copies from individual extraction control replicates were plotted along with each controls’ mean value.

Supp. Fig 5: <u>RT-qPCR Limit of detection.</u> The inset figure shows RT-qPCR generated Ct values for the two-fold dilution series of IDT cDNA copies. The IDT dilution series are 200 (●), 100 (●), 50 (●), 25 (●), 10 (●), 5 (●), 1 (●), and no template control (●; NTC; negative control) (n=12). All replicates were positive when IDT was at higher concentrations (200 to 25 copies); however, 2, 5, and 12 replicates were negative for the IDT at 10, 5, and 1 copies, respectively. Of these 12 replicates per IDT dilution, three representative amplification plots per IDT dilution are presented as normalized reporter value (Rn) by amplification cycle number (Cycle). Color coded labeled arrows point to each triplicate plot.

Supp. Fig 6: <u>Quantification and Comparison of AccuPlex and NATrol Viral Particles</u> (A) AccuPlex and (B) NATrol. RT-qPCR extraction replicates are given in x-axis, and manufacturers’ reported copy numbers are demarcated by red lines. (C) Values reported in the table include particle numbers reported by the manufacturers and RNA copy numbers determined by dd-PCR method and RT-qPCR assays. The fold-increase by RT-qPCR using IDT viral fragments as standard curve is depicted.

Supp. Table 1: <u>Quantification and Comparison of Accuplex and NATrol Viral Particles by direct droplet qPCR</u> Multiple extraction methods were performed, including direct extraction and a variety of RNA extraction kits, followed by ddPCR to determine the precise quantity of N1 and N2 gene copies present in both Accuplex and NATrol viral particles.

Data Set-1: <u>Independent Validation Report of E2E Protocol by a Second laboratory.</u>

## References

1. Kashir J, Yaqinuddin A. 2020. Loop mediated isothermal amplification (LAMP) assays as a rapid diagnostic for COVID-19. Med Hypotheses 141:109786–109786.

2. Ru H, Yang E, Zou K. 2020. What do we learn from SARS-CoV-1 to SARS-CoV-2: Evidence from global stock markets. Available at SSRN 3569330.

3. de Oliveira Araújo FJ, de Lima LSA, Cidade PIM, Nobre CB, Neto MLR. 2020. Impact of Sars-Cov-2 And its reverberation in global higher education and mental health. Psychiatry Res:112977.

4. Han Q, Lin Q, Ni Z, You L. 2020. Uncertainties about the transmission routes of 2019 novel coronavirus. Influenza Other Respi Viruses 14:470–471.

5. West R, Michie S, Rubin GJ, Amlôt R. 2020. Applying principles of behaviour change to reduce SARS-CoV-2 transmission. Nature Human Behaviour: 1-9.

6. Van Doremalen N, Bushmaker T, Morris DH, Holbrook MG, Gamble A, Williamson BN, Tamin A, Harcourt JL, Thornburg NJ, Gerber SI. 2020. Aerosol and surface stability of SARS-CoV-2 as compared with SARS-CoV-1. N Engl J Med 382:1564–1567.

7. Jiang F-C, Jiang X-L, Wang Z-G, Meng Z-H, Shao S-F, Anderson B, Ma M-J. 2020. Detection of Severe Acute Respiratory Syndrome Coronavirus 2 RNA on Surfaces in Quarantine Rooms. Emerging Infectious Disease journal 26.

8. Kampf G, Todt D, Pfaender S, Steinmann E. 2020. Persistence of coronaviruses on inanimate surfaces and their inactivation with biocidal agents. J Hosp Infect 104:246–251.

9. Singhal T. 2020. A review of coronavirus disease-2019 (COVID-19). The Indian Journal of Pediatrics:1-6.

10. Goldman E. 2020. Exaggerated risk of transmission of COVID-19 by fomites. Lancet Infect Dis doi:10.1016/S1473-3099(20)30561-2.

11. Anderson RM, Fraser C, Ghani AC, Donnelly CA, Riley S, Ferguson NM, Leung GM, Lam TH, Hedley AJ. 2004. Epidemiology, transmission dynamics and control of SARS: the 2002-2003 epidemic. Philos Trans R Soc Lond B Biol Sci 359:1091–105.

12. Danko DC, Bezdan D, Afshinnekoo E, Ahsanuddin S, Alicea J, Bhattacharya C, Bhattacharyya M, Blekhman R, Butler DJ, Castro-Nallar E, Cañas AM, Chatziefthimiou AD, Chng KR, Coil DA, Court DS, Crawford RW, Desnues C, Dias-Neto E, Donnellan D, Dybwad M, Eisen JA, Elhaik E, Ercolini D, De Filippis F, Frolova A, Graf AB, Green DC, Lee PKH, Hecht J, Hernandez M, Jang S, Kahles A, Karasikov M, Knights K, Kyrpides NC, Ljungdahl P, Lyons A, Mason-Buck G, McGrath K, Mongodin EF, Mustafa H, Mutai B, Nagarajan N, Neches RY, Ng A, Nieto-Caballero M, Nikolayeva O, Nikolayeva T, Noushmehr H, Oliveira M, et al. 2019. Global Genetic Cartography of Urban Metagenomes and Anti-Microbial Resistance. bioRxiv doi:https://doi.org/10.1101/724526.

13. Kwan K, Cooper M, La Duc MT, Vaishampayan P, Stam C, Benardini JN, Scalzi G, Moissl-Eichinger C, Venkateswaran K. 2011. Evaluation of procedures for the collection, processing, and analysis of biomolecules from low-biomass surfaces. Appl Environ Microbiol 77:2943–2953.

14. Liu H, Gan Y, Yang B, Weng H, Huang C, Yang D, Lei P, Shen G. 2012. Performance evaluation of the Maxwell 16 System for extraction of influenza virus RNA from diverse samples. PLoS One 7:e48094.

15. Dimke H, Larsen SL, Skov MN, Larsen H, Hartmeyer GN, Moeller JB. 2020. Phenol-chloroform-based RNA purification for detection of SARS-CoV-2 by RT-qPCR: comparison with automated systems. medRxiv doi:10.1101/2020.05.26.20099440:2020.05.26.20099440.

16. Wang Y, Kang H, Liu X, Tong Z. 2020. Combination of RT-qPCR testing and clinical features for diagnosis of COVID-19 facilitates management of SARS-CoV-2 outbreak. J Med Virol 92:538–539.

17. Kim D, Lee JY, Yang JS, Kim JW, Kim VN, Chang H. 2020. The Architecture of SARS-CoV-2 Transcriptome. Cell 181:914–921 e10.

18. Meyer Kate D, Saletore Y, Zumbo P, Elemento O, Mason Christopher E, Jaffrey Samie R. 2012. Comprehensive Analysis of mRNA Methylation Reveals Enrichment in 3’ UTRs and near Stop Codons. Cell 149:1635–1646.

19. Viehweger A, Krautwurst S, Lamkiewicz K, Madhugiri R, Ziebuhr J, Hölzer M, Marz M. 2019. Direct RNA nanopore sequencing of full-length coronavirus genomes provides novel insights into structural variants and enables modification analysis. Genome Res 29:1545–1554.

20. Hung SS, Meissner B, Chavez EA, Ben-Neriah S, Ennishi D, Jones MR, Shulha HP, Chan FC, Boyle M, Kridel R, Gascoyne RD, Mungall AJ, Marra MA, Scott DW, Connors JM, Steidl C. 2018. Assessment of Capture and Amplicon-Based Approaches for the Development of a Targeted Next-Generation Sequencing Pipeline to Personalize Lymphoma Management. The Journal of Molecular Diagnostics 20:203–214.

21. Lu R, Wang J, Li M, Wang Y, Dong J, Cai W. 2020. SARS-CoV-2 detection using digital PCR for COVID-19 diagnosis, treatment monitoring and criteria for discharge. medRxiv.

22. Parida M, Posadas G, Inoue S, Hasebe F, Morita K. 2004. Real-time reverse transcription loop-mediated isothermal amplification for rapid detection of West Nile virus. J Clin Microbiol 42:257–263.

23. Parida M, Santhosh S, Dash P, Tripathi N, Lakshmi V, Mamidi N, Shrivastva A, Gupta N, Saxena P, Babu JP. 2007. Rapid and real-time detection of Chikungunya virus by reverse transcription loop-mediated isothermal amplification assay. J Clin Microbiol 45:351–357.

24. Martin-Latil S, Hennechart-Collette C, Guillier L, Perelle S. 2012. Comparison of two extraction methods for the detection of hepatitis A virus in semi-dried tomatoes and murine norovirus as a process control by duplex RT-qPCR. Food Microbiol 31:246–253.

25. Dupont-Rouzeyrol M, Biron A, O’Connor O, Huguon E, Descloux E. 2016. Infectious Zika viral particles in breastmilk. The Lancet 387:1051.

26. de Crom SCM, Obihara CC, de Moor RA, Veldkamp EJM, van Furth AM, Rossen JWA. 2013. Prospective comparison of the detection rates of human enterovirus and parechovirus RT-qPCR and viral culture in different pediatric specimens. Journal of clinical virology: the official publication of the Pan American Society for Clinical Virology 58:449–454.

27. Leung NH, Chu DK, Shiu EY, Chan K-H, McDevitt JJ, Hau BJ, Yen H-L, Li Y, Ip DK, Peiris JM. 2020. Respiratory virus shedding in exhaled breath and efficacy of face masks. Nat Med 26:676–680.

28. Boone SA, Gerba CP. 2007. Significance of fomites in the spread of respiratory and enteric viral disease. Appl Environ Microbiol 73:1687–1696.

29. Cai J, Sun W, Huang J, Gamber M, Wu J, He G. 2020. Indirect virus transmission in cluster of COVID-19 cases, Wenzhou, China, 2020.

30. Liu K, Chen Y, Lin R, Han K. 2020. Clinical features of COVID-19 in elderly patients: A comparison with young and middle-aged patients. J Infect.

31. MacIntyre CR. 2014. The discrepant epidemiology of Middle East respiratory syndrome coronavirus (MERS-CoV). Environ Syst Decis 34:383–390.

32. Elimelech M, Xiaohua Z, Childress AE, Seungkwan H. 1997. Role of membrane surface morphology in colloidal fouling of cellulose acetate and composite aromatic polyamide reverse osmosis membranes. J Membr Sci 127:101–109.

33. Yamagishi T. 2020. Environmental sampling for severe acute respiratory syndrome coronavirus 2 (SARS-CoV-2) during a coronavirus disease (COVID-19) outbreak aboard a commercial cruise ship. medRxiv doi:10.1101/2020.05.02.20088567:2020.05.02.20088567.

34. Bruce EA, Huang M-L, Perchetti GA, Tighe S, Laaguiby P, Hoffman JJ, Gerrard DL, Nalla AK, Wei Y, Greninger AL, Diehl SA, Shirley DJ, Leonard DGB, Huston CD, Kirkpatrick BD, Dragon JA, Crothers JW, Jerome KR, Botten JW. 2020. DIRECT RT-qPCR DETECTION OF SARS-CoV-2 RNA FROM PATIENT NASOPHARYNGEAL SWABS WITHOUT AN RNA EXTRACTION STEP. bioRxiv doi:10.1101/2020.03.20.001008:2020.03.20.001008.

35. Bargoma E, La Duc MT, Kwan K, Vaishampayan P, Venkateswaran K. 2013. Differential recovery of phylogenetically disparate microbes from spacecraft-qualified metal surfaces. Astrobiology 13:189–202.

36. Ong SWX, Tan YK, Chia PY, Lee TH, Ng OT, Wong MSY, Marimuthu K. 2020. Air, Surface Environmental, and Personal Protective Equipment Contamination by Severe Acute Respiratory Syndrome Coronavirus 2 (SARS-CoV-2) From a Symptomatic Patient. JAMA 323:1610–1612.

37. Kwan K, Cooper M, La Duc MT, Vaishampayan P, Stam C, Benardini JN, Scalzi G, Moissl-Eichinger C, Venkateswaran K. 2011. Evaluation of procedures for the collection, processing, and analysis of biomolecules from low-biomass surfaces. Appl Environ Microbiol 77:2943–53.

38. La Duc MT, Osman S, Venkateswaran K. 2009. Comparative analysis of methods for the purification of DNA from low-biomass samples based on total yield and conserved microbial diversity. Journal of Rapid Methods & Automation in Microbiology 17:350-368.

39. Danko DC, Bezdan D, Afshinnekoo E, Ahsanuddin S, Alicea J, Bhattacharya C, Bhattacharyya M, Blekhman R, Butler DJ, Castro-Nallar E, Cañas AM, Chatziefthimiou AD, Chng KR, Coil DA, Court DS, Crawford RW, Desnues C, Dias-Neto E, Donnellan D, Dybwad M, Eisen JA, Elhaik E, Ercolini D, De Filippis F, Frolova A, Graf AB, Green DC, Lee PKH, Hecht J, Hernandez M, Jang S, Kahles A, Karasikov M, Knights K, Kyrpides NC, Ljungdahl P, Lyons A, Mason-Buck G, McGrath K, Mongodin EF, Mustafa H, Mutai B, Nagarajan N, Neches RY, Ng A, Nieto-Caballero M, Nikolayeva O, Nikolayeva T, Noushmehr H, Oliveira M, et al. 2019. Global Genetic Cartography of Urban Metagenomes and Anti-Microbial Resistance. bioRxiv doi:10.1101/724526:724526.

40. Corman VM, Landt O, Kaiser M, Molenkamp R, Meijer A, Chu DK, Bleicker T, Brünink S, Schneider J, Schmidt ML, Mulders DG, Haagmans BL, van der Veer B, van den Brink S, Wijsman L, Goderski G, Romette J-L, Ellis J, Zambon M, Peiris M, Goossens H, Reusken C, Koopmans MP, Drosten C. 2020. Detection of 2019 novel coronavirus (2019-nCoV) by real-time RT-PCR. Euro surveillance: bulletin Europeen sur les maladies transmissibles = European communicable disease bulletin 25:2000045.

41. Butler DJ, Mozsary C, Meydan C, Danko D, Foox J, Rosiene J, Shaiber A, Afshinnekoo E, MacKay M, Sedlazeck FJ, Ivanov NA, Sierra M, Pohle D, Zietz M, Gisladottir U, Ramlall V, Westover CD, Ryon K, Young B, Bhattacharya C, Ruggiero P, Langhorst BW, Tanner N, Gawrys J, Meleshko D, Xu D, Steel PAD, Shemesh AJ, Xiang J, Thierry-Mieg J, Thierry-Mieg D, Schwartz RE, Iftner A, Bezdan D, Sipley J, Cong L, Craney A, Velu P, Melnick AM, Hajirasouliha I, Horner SM, Iftner T, Salvatore M, Loda M, Westblade LF, Cushing M, Levy S, Wu S, Tatonetti N, Imielinski M, et al. 2020. Shotgun Transcriptome and Isothermal Profiling of SARS-CoV-2 Infection Reveals Unique Host Responses, Viral Diversification, and Drug Interactions. bioRxiv doi:10.1101/2020.04.20.048066:2020.04.20.048066.

42. McEnerney B. 2018. General Cleaning of Materials. Jet Propulsion Laboratoy, California Institute of Technology, Pasadena, CA, USA.

